# Integrating Serum Biomarkers and Genetic Polymorphisms for Alzheimer’s Disease Risk Screening

**DOI:** 10.1101/2025.06.17.25329820

**Authors:** Zhizhen Li, Zhaohai Feng, Song He, Huaping Tang, Cai Zhao, Jinfeng Liu, Yao Su, Jiayang Duan, Xue Tian, Yong Yan, Xiaojing Shi, Xueling Bi

## Abstract

**Objective:** Analyze the differences in the detection results of Alzheimer’s disease (AD) serum biomarkers and the expression of APOE and SLCO1B1 genotypes, then construct an AD diagnostic model integrating serum biomarkers with APOE and SLCO1B1 genotypes to enhance the sensitivity and specificity of AD screening.

**Methods:** Serum samples from 127 participants (41 AD, 39 cognitive impairments, 47 controls) were analyzed for neurofilament light chain (NF), Aβ42/Aβ40 ratio, p-TAU231, and p-TAU217 via ELISA, combined with APOE/SLCO1B1 genotyping. A nomogram model was constructed using logistic regression and validated by ROC-AUC and Hosmer-Lemeshow tests.

**Results:** The AD group showed elevated NF (*p*<0.01) and p-TAU231 (*p*<0.05), reduced Aβ42/Aβ40 (*p*<0.05). APOE ε4 homozygotes exhibited the highest p-TAU181 (*p*<0.001), while SLCO1B1*1b/*1b correlated with increased Aβ42 (*p*<0.05). The biomarker model achieved AUC=0.8365, improving to 0.8832 after genetic integration (calibration *p*>0.05).

**Conclusion:** This study pioneers an AD diagnostic model combining serum p-TAU231, NF, and SLCO1B1 polymorphisms, surpassing single-marker limitations (AUC>0.88). It provides a high-accuracy tool for non-invasive screening and genetic stratification, demonstrating substantial clinical potential.

## Introduction

Alzheimer’s Disease (AD) is a neurodegenerative disorder characterized by progressive cognitive decline and neuronal loss, affecting over 55 million individuals globally, with projections estimating a rise to 152 million by 2050 [1]. Its pathological hallmarks include senile plaques formed by β-amyloid (Aβ) deposition, neurofibrillary tangles resulting from hyperphosphorylated tau protein, accompanied by neuroinflammation and synaptic dysfunction [2,3]. Despite significant advances in understanding AD pathophysiology, early diagnosis remains challenging: cerebrospinal fluid (CSF) biomarker assays and positron emission tomography (PET) are costly and invasive, while clinical symptoms typically manifest only after irreversible neuronal damage has occurred [4]. Consequently, the development of highly sensitive, non-invasive peripheral biomarkers and diagnostic models integrating genetic risk profiles has emerged as a critical research priority.

In recent years, blood-based biomarkers have garnered attention due to their accessibility and reproducibility. Neurofilament light chain (NF), a marker of axonal injury, is significantly elevated in the serum of AD patients; however, its low specificity overlaps with other neurodegenerative disorders, such as frontotemporal dementia [5,6]. Reduced Aβ42/Aβ40 ratios and elevated phosphorylated tau (p-tau) protein levels, particularly p-tau231 and p-tau217, have been strongly associated with AD pathology, with their stability in plasma rendering them promising diagnostic tools [7,8]. Nevertheless, the diagnostic efficacy of individual biomarkers remains limited, while combining multiple biomarkers may lead to improved diagnostic performance, and interindividual genetic heterogeneity may influence biomarker expression [9]. For instance, the apolipoprotein E (APOE) ε4 allele, the strongest genetic risk factor for AD, correlates with impaired Aβ clearance and accelerated tau pathology progression [10], while SLCO1B1 gene polymorphisms may indirectly modulate AD risk through lipid metabolism regulation and statin pharmacodynamics [11]. These findings underscore the potential of integrating multidimensional biomarkers and genetic data to overcome current diagnostic limitations.

Existing studies often examine single biomarkers or genetic factors in isolation, neglecting their potential interactions. Although the ATN (Amyloid-Tau-Neurodegeneration) framework proposes a diagnostic strategy combining Aβ, tau, and neurodegeneration markers [12,13], the incorporation of genetic variants into this system remains undefined. Addressing these gaps, this study analyzes serum levels of NF, Aβ42/Aβ40, p-tau231, and p-tau217 in AD patients, individuals with Cognitive Impairment (CI), and healthy controls, combined with APOE and SLCO1B1 genotyping. Our aim is to evaluate the diagnostic utility of these biomarkers for AD and to construct a risk-prediction nomogram model based on their profiles.

## Materials and Methods

### Study Participants and Design

Serum samples were collected from 127 hospitalized patients in the Department of Neurology and Geriatrics at Maanshan People’s Hospital between 01/02/2024 and 31/01/2025. Participants were categorized into three groups based on diagnostic criteria: the AD group (n=41), the CI group (n=39), and the cognitively normal control group (n=47). The diagnosis of AD was based on the McKhann criteria [14], while the diagnosis of CI followed the Peterson criteria [15]. Clinical data were recorded, and serum levels of NF, Aβ40, Aβ42 (to calculate the Aβ42/Aβ40 ratio), p-tau181, p-tau217, p-tau231, as well as SLCO1B1 and APOE genotypes, were retrospectively analyzed. Intergroup differences in biomarker expression and their associations with APOE and SLCO1B1 polymorphisms were evaluated. Diagnostic performance of these biomarkers for AD was assessed, and a risk-prediction nomogram model was developed and validated. This research was a retrospective investigation. Patient identifiers were removed during data extraction, with only gender, age, clinical diagnoses, and laboratory test data collected. This study was approved by the Ethics Committee of Maanshan People’s Hospital (Approval No.: ERMPH 2025 No 02-07 V1.0 2024.1.30), and the requirement for informed consent was waived due to the retrospective nature of the data analysis.

### Assay Reagents and Equipments

Serum levels of NF, Aβ40, Aβ42, p-tau181, p-tau217, and p-tau231 were quantified using enzyme-linked immunosorbent assay (ELISA). The NF ELISA kit was supplied by Jingmei Biotechnology Co., Ltd., while other reagents were provided by Shanghai Jianglai Biotechnology Co., Ltd. Assays were performed on a fully automated ELISA workstation (Model: ADC ELISA 1100; Yantai Aideng Biotechnology Co., Ltd.). Genotyping of APOE and SLCO1B1 was conducted via polymerase chain reaction (PCR). Nucleic acid extraction kits were obtained from Jiangsu Kangwei Century Biotechnology Co., Ltd., and PCR reagents were sourced from Wuhan Youzhiyou Medical Technology Co., Ltd. PCR analysis was performed using an automated medical PCR system (Xi’an Tianlong Technology Co., Ltd.). All procedures strictly followed manufacturer protocols and equipment specifications.

### Statistical Analysis

Data were organized using Microsoft Excel 2019 and analyzed with SPSS 20, R (v4.4.3), and STATA 15. Categorical variables were compared using chi-square tests, while quantitative data were analyzed in R with appropriate statistical methods (stats and car packages). As most data deviated from normality, intergroup differences were assessed via the *Kruskal-Wallis* test, followed by *Dunn’s* post-hoc test for pairwise comparisons. Diagnostic performance was evaluated using receiver operating characteristic (ROC) curves and area under the curve (AUC) calculations. Data visualization was performed using ggplot2 in R. Statistical significance was defined as *p*<0.05. The nomogram model was constructed using R (rms package) and STATA, with validation metrics including ROC analysis for discriminative ability and the *Hosmer-Lemeshow* test for calibration (*p*>0.05 indicating good agreement between predicted and observed values). The significance of differences in the AUC-ROC was assessed using DeLong’s test, with *p*<0.05 indicating statistical significance.

## Results

### Baseline Demographic Characteristics

Among the study participants, the AD group included 13 males (31.71%), the CI group had 14 males (35.90%), and the cognitively normal control group comprised 19 males (40.43%). No statistically significant differences in gender distribution were observed across groups (χ²=0.723, *p*=0.697). As shown in Table 1, age distributions also did not differ significantly among the three groups (*p*>0.05), confirming baseline comparability.

**Table 1.**
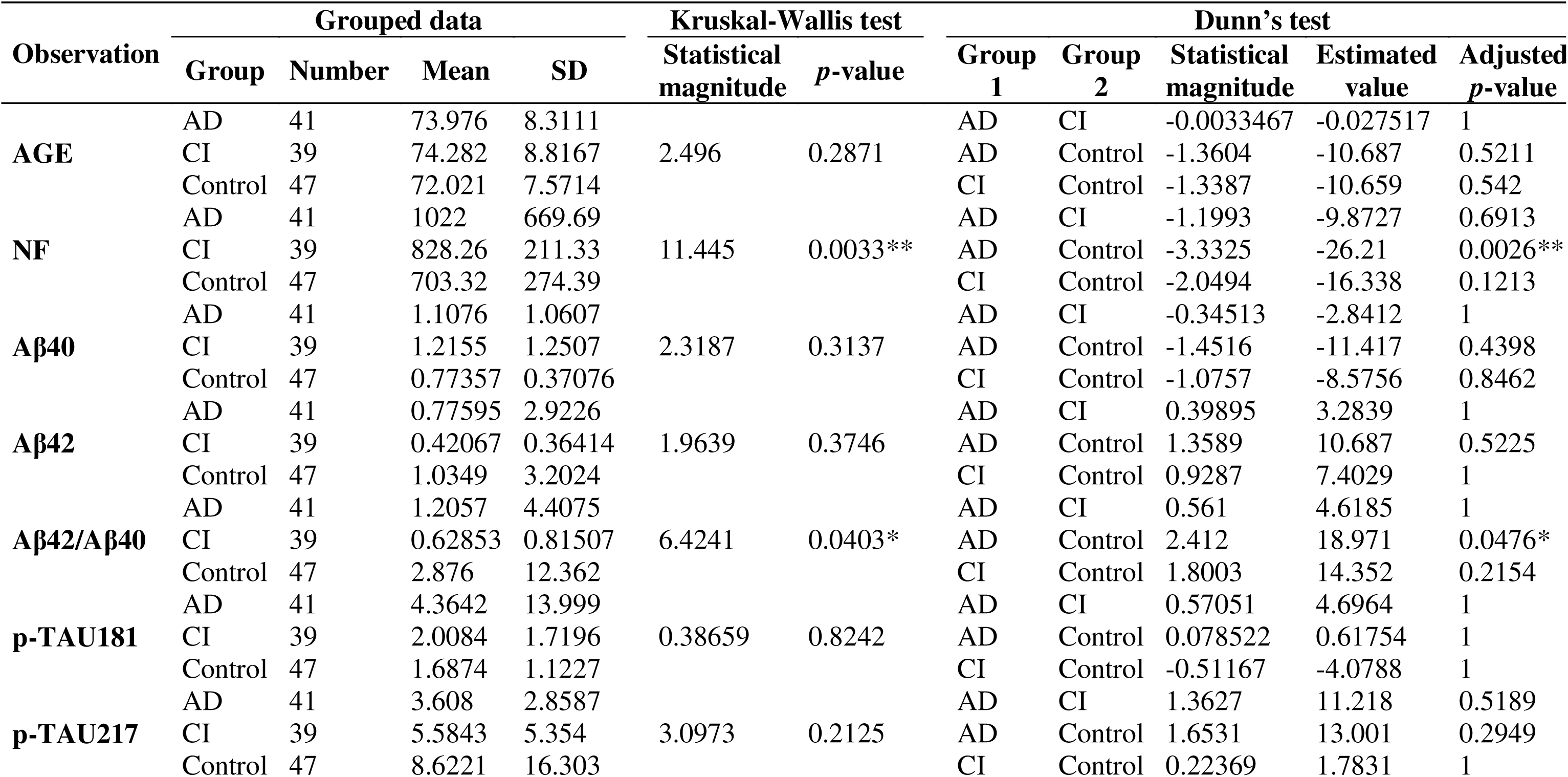
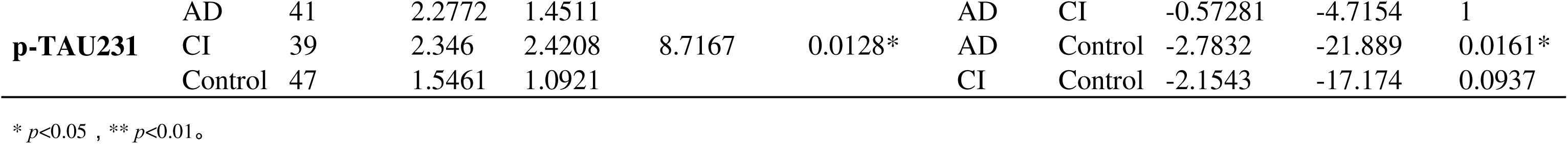
Descriptive Statistics and Intergroup Comparative Analysis of Biomarkers.

| Observation | Grouped data |  |  |  | Kruskal-Wallis test |  | Dunn's test |  |  |  |  |
| --- | --- | --- | --- | --- | --- | --- | --- | --- | --- | --- | --- |
|  | Group | Number | Mean | SD | Statistical magnitude | *p*-value | Group 1 | Group 2 | Statistical magnitude | Estimated value | Adjusted *p*-value |
| AGE | AD | 41 | 73.976 | 8.3111 | 2.496 | 0.2871 | AD | CI | -0.0033467 | -0.027517 | 1 |
|  | CI | 39 | 74.282 | 8.8167 |  |  | AD | Control | -1.3604 | -10.687 | 0.5211 |
|  | Control | 47 | 72.021 | 7.5714 |  |  | CI | Control | -1.3387 | -10.659 | 0.542 |
| NF | AD | 41 | 1022 | 669.69 | 11.445 | 0.0033** | AD | CI | -1.1993 | -9.8727 | 0.6913 |
|  | CI | 39 | 828.26 | 211.33 |  |  | AD | Control | -3.3325 | -26.21 | 0.0026** |
|  | Control | 47 | 703.32 | 274.39 |  |  | CI | Control | -2.0494 | -16.338 | 0.1213 |
| Aβ40 | AD | 41 | 1.1076 | 1.0607 | 2.3187 | 0.3137 | AD | CI | -0.34513 | -2.8412 | 1 |
|  | CI | 39 | 1.2155 | 1.2507 |  |  | AD | Control | -1.4516 | -11.417 | 0.4398 |
|  | Control | 47 | 0.77357 | 0.37076 |  |  | CI | Control | -1.0757 | -8.5756 | 0.8462 |
| Aβ42 | AD | 41 | 0.77595 | 2.9226 | 1.9639 | 0.3746 | AD | CI | 0.39895 | 3.2839 | 1 |
|  | CI | 39 | 0.42067 | 0.36414 |  |  | AD | Control | 1.3589 | 10.687 | 0.5225 |
|  | Control | 47 | 1.0349 | 3.2024 |  |  | CI | Control | 0.9287 | 7.4029 | 1 |
| Aβ42/Aβ40 | AD | 41 | 1.2057 | 4.4075 | 6.4241 | 0.0403* | AD | CI | 0.561 | 4.6185 | 1 |
|  | CI | 39 | 0.62853 | 0.81507 |  |  | AD | Control | 2.412 | 18.971 | 0.0476* |
|  | Control | 47 | 2.876 | 12.362 |  |  | CI | Control | 1.8003 | 14.352 | 0.2154 |
| p-TAU181 | AD | 41 | 4.3642 | 13.999 | 0.38659 | 0.8242 | AD | CI | 0.57051 | 4.6964 | 1 |
|  | CI | 39 | 2.0084 | 1.7196 |  |  | AD | Control | 0.078522 | 0.61754 | 1 |
|  | Control | 47 | 1.6874 | 1.1227 |  |  | CI | Control | -0.51167 | -4.0788 | 1 |
| p-TAU217 | AD | 41 | 3.608 | 2.8587 | 3.0973 | 0.2125 | AD | CI | 1.3627 | 11.218 | 0.5189 |
|  | CI | 39 | 5.5843 | 5.354 |  |  | AD | Control | 1.6531 | 13.001 | 0.2949 |
|  | Control | 47 | 8.6221 | 16.303 |  |  | CI | Control | 0.22369 | 1.7831 | 1 |
| **p-TAU231** | AD | 41 | 2.2772 | 1.4511 | 8.7167 | 0.0128* | AD | CI | -0.57281 | -4.7154 | 1 |
|  | CI | 39 | 2.346 | 2.4208 |  |  | AD | Control | -2.7832 | -21.889 | 0.0161* |
|  | Control | 47 | 1.5461 | 1.0921 |  |  | CI | Control | -2.1543 | -17.174 | 0.0937 |

\* $p<0.05$ , \*\* $p<0.01$ 。

### Intergroup Differences in Biomarker Levels

Biomarker data and intergroup comparisons are summarized in Table 1. Significant differences (*p*<0.05) were observed across groups in serum levels of NF, Aβ42/Aβ40, and p-tau231. NF levels exhibited an increasing trend from the cognitively normal control group to the CI group and AD group, with pairwise comparisons confirming significantly higher NF in the AD group versus controls (*p*<0.01). The Aβ42/Aβ40 ratio was significantly lower in the AD group compared to controls (*p*<0.05), while p-tau231 levels were markedly elevated in the AD group (*p*<0.05). Although p-tau217 levels did not differ significantly across groups, a decreasing trend was noted from controls to the CI and AD groups.

### Diagnostic Performance of Biomarkers for AD

Receiver operating characteristic (ROC) curves were generated to evaluate the diagnostic utility of biomarkers for AD (Fig 1). NF demonstrated the highest discriminative power (AUC=0.701), followed by p-tau231 (AUC=0.668), Aβ42/Aβ40 (AUC=0.651), and p-tau217 (AUC=0.604).

**Fig 1.**
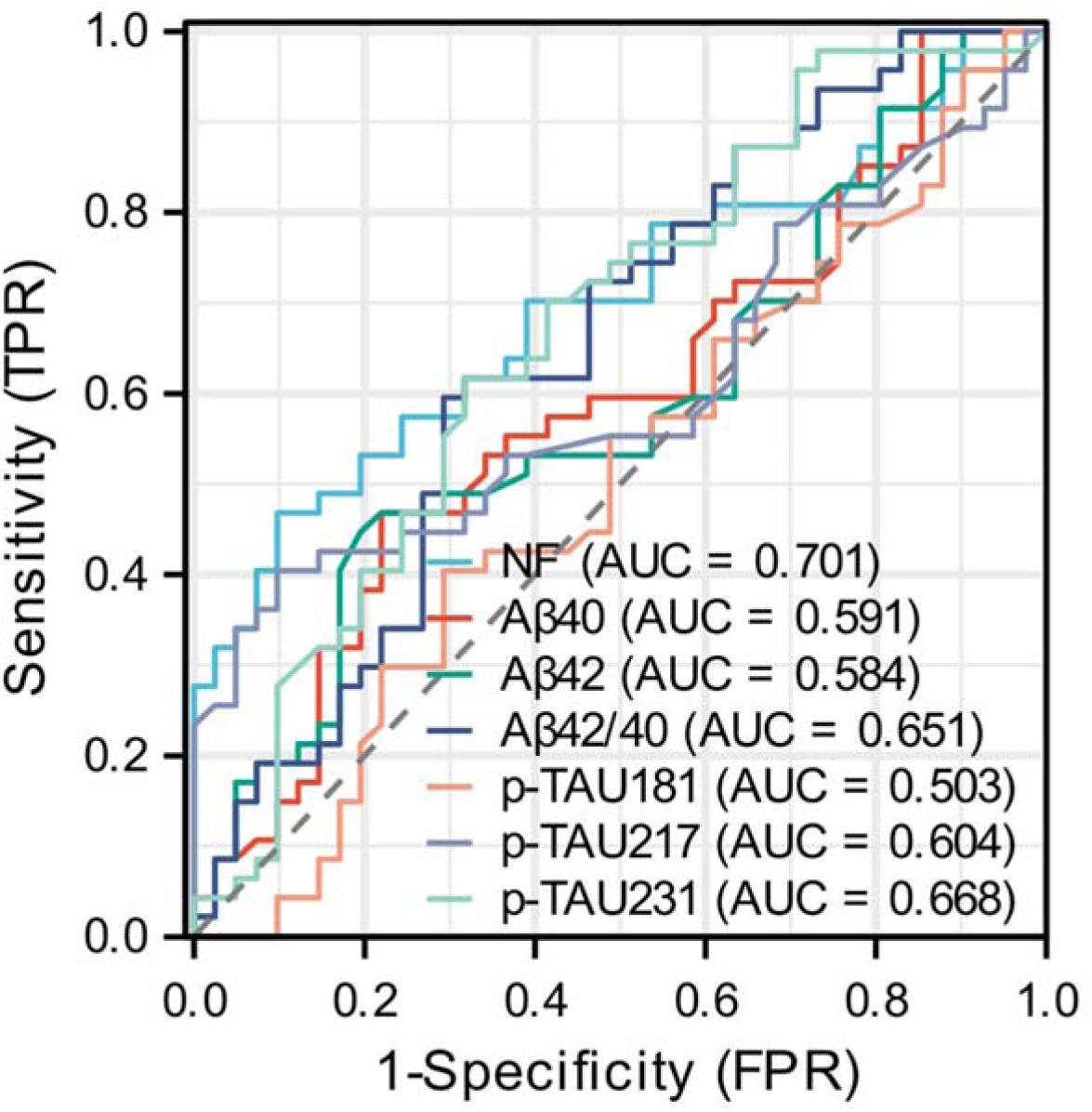
ROC Curve Analysis of Serum Biomarkers for the Diagnosis of Alzheimer’s Disease.

### Association of Biomarkers with APOE and SLCO1B1 Genotypes

Genetic analysis revealed significant associations between biomarker levels and APOE/SLCO1B1 polymorphisms.

For APOE genotypes: NF levels were significantly higher in the ε2/ε4 group compared to ε3/ε3 carriers (*p*=0.0049). Aβ40 levels were elevated in the ε2/ε4 group versus ε3/ε4 carriers (*p*=0.0447). p-tau181 levels were markedly higher in ε4/ε4 carriers than in ε3/ε4 (*p*=0.0001), ε2/ε4 (*p*=0.0447), and ε2/ε3 (*p*=0.0464) groups. ε3/ε3 carriers also showed higher p-tau181 levels than ε3/ε4 carriers (*p*=7.81×10). p-tau217 levels were significantly elevated in ε3/ε3 versus ε3/ε4 carriers (*p*=0.0399) (Fig 2(a)).

**Fig 2.**
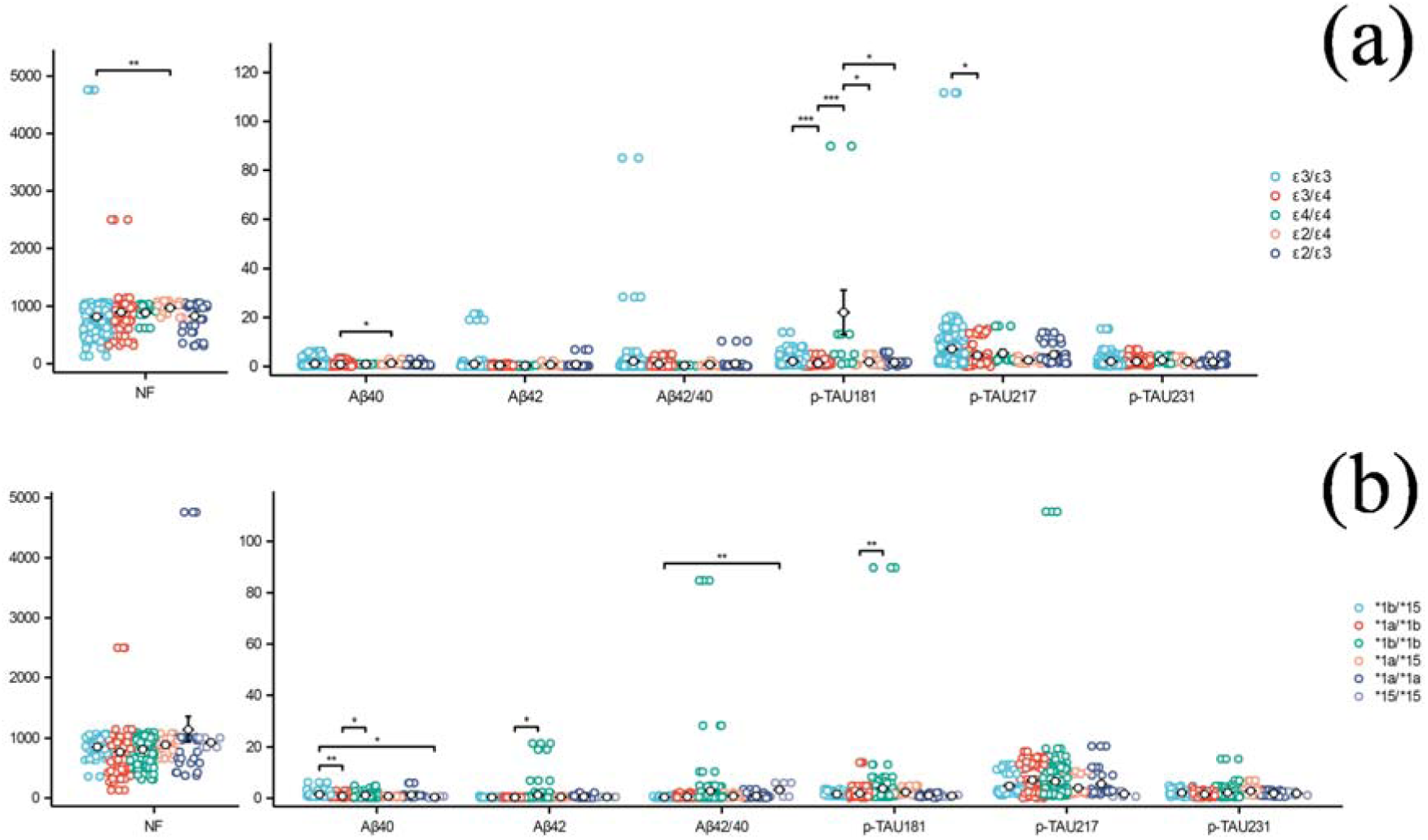
Association of AD Biomarkers with APOE and SLCO1B1 Genotypes. (a). Association of Alzheimer’s Disease-Associated Serum Biomarkers with APOE Genotypes. (b). Association of Alzheimer’s Disease-Associated Serum Biomarkers with SLCO1B1 Genotypes. * p<0.05, ** p<0.01, ***p<0.001_。_

For SLCO1B1 genotypes: Aβ40 levels were higher in *1b/*15 carriers compared to *1a/*1b (p=0.0015) and *15/*15 (p=0.0155) groups, with *1b/*1b carriers also showing elevated Aβ40 versus *1a/*1b (*p*=0.0102). Aβ42 levels were increased in *1b/*1b versus *1a/*1b carriers (*p*=0.0112). The Aβ42/Aβ40 ratio was higher in *15/*15 versus *1b/*15 carriers (*p*=0.0043). p-tau181 levels were significantly lower in *1a/*1b carriers compared to *1b/*1b (*p*=0.005) and *1a/*15 (*p*=0.0064) groups, with *1a/*1a and *15/*15 carriers also showing reduced levels versus *1b/*1b (*p*=0.0002) and *1a/*15 (*p*=0.0002) groups. p-tau231 levels were lower in *1a/*1b carriers compared to *1a/*15 (*p*=0.0205) and *1b/*15 (*p*=0.0003) groups (Fig 2(b)).

### Construction of the AD Risk-Prediction Nomogram Model

To enhance the diagnostic accuracy for AD risk, we performed logistic regression analysis on the biomarkers and constructed a nomogram model (Fig 3(a)). Discrimination validation via ROC curve analysis revealed an AUC of 0.8365, surpassing the diagnostic performance of individual biomarkers (Fig 3(b)). Calibration validation using the *Hosmer-Lemeshow* test yielded a Pearson χ²=88.73 (*p*=0.2127), indicating good agreement between predicted and observed values. Given the observed associations between APOE/SLCO1B1 genotypes and AD biomarker levels, we further incorporated genetic variants into the nomogram (Fig 3(c)). This integrated model demonstrated increased diagnostic efficacy (*p*=0.0939, Delong’s test), with an AUC-ROC of 0.8832 (Fig 3(d)) and a *Hosmer-Lemeshow* test result of χ²=81.47 (*p*=0.313), confirming robust calibration.

**Fig 3.**
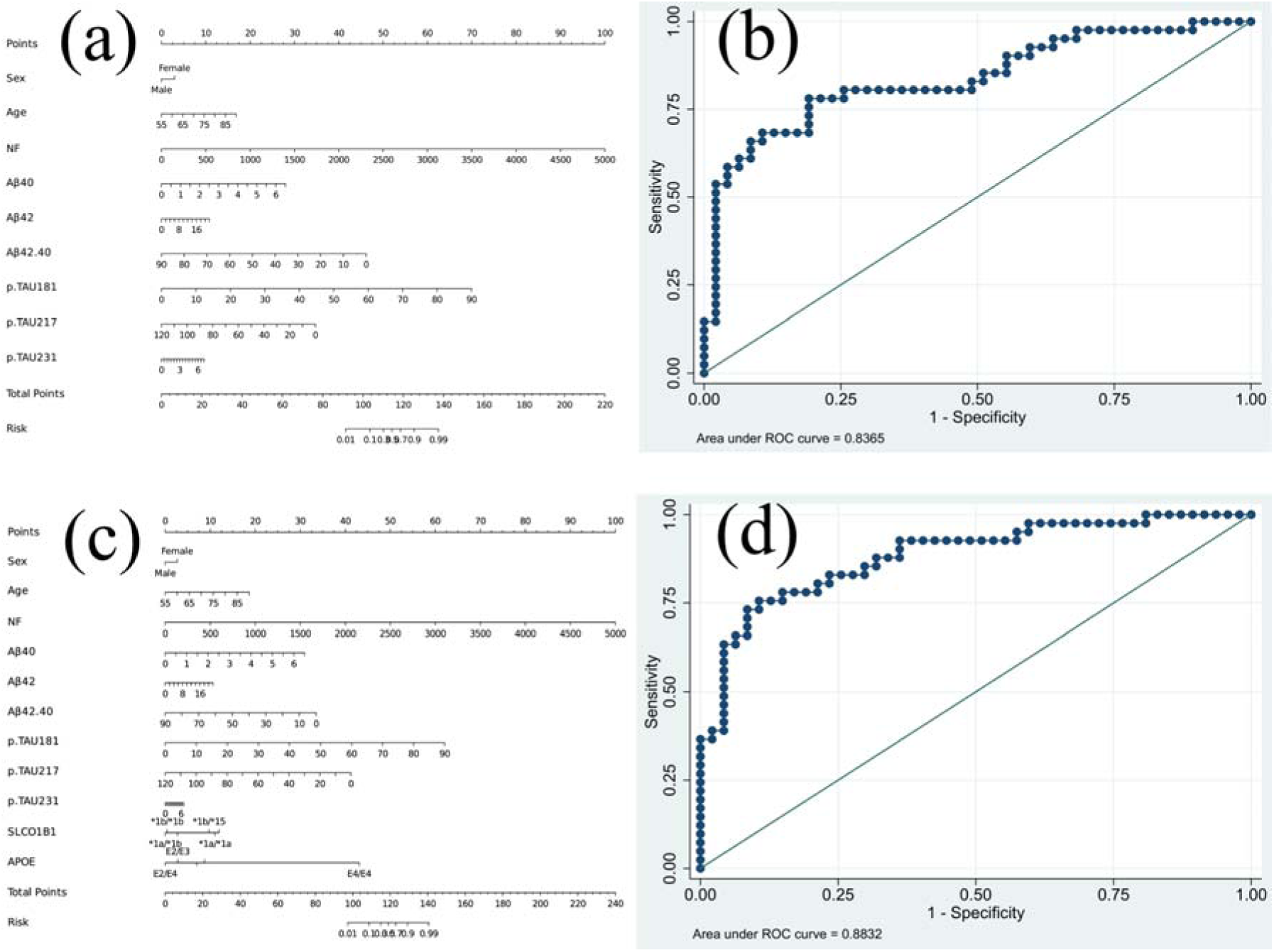
AD Risk-Prediction Nomogram Model. (a). Nomogram for Alzheimer’s Disease Risk Diagnosis Based on Combined Serum Biomarkers. (b). ROC Curve Validation of the Nomogram for Alzheimer’s Disease Risk Diagnosis. (c). Nomogram for Alzheimer’s Disease Risk Diagnosis Incorporating Serum Biomarkers with APOE and SLCO1B1 Genotyping. (d). ROC curve validation of the nomogram for Alzheimer’s disease risk diagnosis after incorporating APOE and SLCO1B1 genotypes.

## Discussion

AD is a neurodegenerative disorder characterized by amyloid-β (Aβ) deposition, aberrant tau phosphorylation, and neuronal degeneration. Recent advancements in biomarker research have provided critical insights into early diagnosis and pathological mechanisms. This study developed an AD risk-prediction model by integrating serum biomarkers (NF, Aβ42/Aβ40 ratio, p-tau231, p-tau217) with APOE and SLCO1B1 polymorphisms.

Our findings revealed significantly elevated NF levels in the AD group compared to cognitively normal controls (p<0.01), with a gradient trend (AD > CI > controls). NF, a marker of axonal injury, reflects progressive neuronal degeneration [5]. Previous studies have correlated NF with neurodegeneration severity in AD CSF [16], and our results further validate its diagnostic potential in serum. However, NF lacks disease specificity, as it is also elevated in other neurodegenerative conditions (e.g., Par-kinson’s disease, multiple sclerosis) [17,18], necessitating integration with other bio-markers to enhance diagnostic specificity.

The reduced Aβ42/Aβ40 ratio in the AD group (p<0.05) aligns with the pathological accumulation and impaired clearance of Aβ42 in AD [2,3]. A low Aβ42/Aβ40 ratio is widely recognized as a core biomarker in both CSF and plasma [7,8]. Additionally, ele-vated p-tau231 levels in the AD group (p<0.05) underscore the role of tau hyperphos-phorylation in AD progression. Notably, while p-tau217 did not show significant inter-group differences, its decreasing trend from controls to the AD group may suggest temporal specificity in phosphorylation dynamics, warranting longitudinal investigation.

NF demonstrated the highest diagnostic sensitivity (AUC=0.701) compared to p-tau231 (0.668) and Aβ42/Aβ40 (0.651). However, the limited discriminative power of single biomarkers highlights the need for multimodal approaches. For instance, the ATN framework advocates combining Aβ, tau, and neurodegeneration markers—a strategy corroborated by our findings.

Our data also revealed findings that are inconsistent with widely accepted evidence, such as: As shown in Table 1, several key biomarkers exhibit counterintuitive patterns. The cognitively impaired (CI) group, rather than the AD group, shows the lowest levels of Aβ42 and the Aβ42/40 ratio, and the highest levels of Aβ40 and p-tau231. This suggests a more severe pathological profile in the CI group compared to the diagnosed AD group, which is biologically unexpected. The trend for p-tau217 (Control > CI > AD) is in direct opposition to a large body of established literature, which consistently demonstrates that plasma p-tau217 is significantly elevated in AD. These discrepancies are likely attributable to the suboptimal sensitivity of the assay methodology we employed. This observation nonetheless provides valuable clinical insight: more advanced and highly sensitive detection platforms, such as SIMOA [19–20], should be prioritized for measuring AD biomarkers.

The APOE ε4 allele remains the strongest genetic risk factor for AD [21]. Our study revealed significantly elevated p-tau181 levels in APOE ε4/ε4 genotype carriers compared to other genotypes (p<0.001), suggesting that ε4 may accelerate AD progression by promoting tau phosphorylation. Additionally, the ε2/ε4 genotype was associated with increased NF levels (p=0.0049), implying a potential neuroprotective role of the ε2 allele in counteracting ε4-mediated toxicity [7]. These findings reinforce the utility of APOE genotyping in AD subtyping and prognostic evaluation.

SLCO1B1, encoding the hepatic organic anion-transporting polypeptide, influences statin metabolism and Aβ clearance through its polymorphisms 9. In our cohort, *1b/*1b carriers exhibited higher Aβ42 levels than *1a/*1b carriers (p=0.0112), while the *15/*15 genotype was associated with an elevated Aβ42/Aβ40 ratio (p=0.0043). These results suggest that SLCO1B1 variants may modulate Aβ homeostasis via lipid metabolism regulation. Furthermore, genotype-dependent differences in p-tau181 and p-tau231 levels (e.g., lower p-tau in *1a/*1b carriers) could inform the development of genotype-guided therapies.

The nomogram model constructed via logistic regression achieved an AUC of 0.8365, significantly outperforming individual biomarkers (e.g., NF: AUC=0.701). The model demonstrated excellent calibration (*Hosmer-Lemeshow* test p>0.05), indicating high concordance between predicted and observed risks. Upon integrating APOE and SLCO1B1 genotypes, the AUC increased to 0.8832, underscoring the critical role of genetic data in AD stratification. Similar strategies, such as the polygenic risk score (PRS) model by Kunkle *et al*. [11], validate the superiority of combined genetic-biomarker approaches.

This model provides a novel tool for early AD screening and risk stratification, particularly in resource-limited settings where CSF or PET imaging is inaccessible. However, its clinical translation requires addressing several limitations. Firstly, sample size constraints: The relatively small AD cohort (n=41) necessitates validation in larger populations. Secondly, unaccounted clinical variables: Cognitive scores and neuroimaging data were not included. Thirdly, population genetic heterogeneity: Generalizability across ethnic groups remains unverified.

As a cross-sectional study, our design precludes causal inferences between biomarkers and AD progression. Future longitudinal studies should delineate dynamic changes in NF, p-tau231, and other biomarkers across disease stages. Additionally, the interplay between SLCO1B1 polymorphisms and statin pharmacodynamics warrants exploration, potentially guiding personalized treatment for AD patients with comorbid hyperlipidemia.

## Data Availability

The data underlying the results presented in the study are available from the Corresponding author.

## Abbreviations

Aβ: β-amyloid
AD: Alzheimer’s disease
APOE: apolipoprotein E
ATN: Amyloid-Tau-Neurodegeneration
AUC: Area Under the Curve
CI: Cognitive Impairment
CSF: cerebrospinal fluid
ELISA: Enzyme-linked immunosorbent assay
NF: Neurofilament light chain
p-tau: phosphorylated tau
PCR: polymerase chain reaction
PET: positron emission tomography
ROC: receiver operating characteristic
SLCO1B1: Solute carrier organic anion transporter family member 1B1

## Acknowledgements

We would like to express our gratitude to all the members of our research team for their contributions in experimental testing and data management.

